# Multiscale predictability of Cutaneous Leishmaniasis in Morocco and Tunisia through the AMO-NAO coupling and its modulation of regional rainfall

**DOI:** 10.1101/2023.12.14.23299949

**Authors:** Adrià San-José, Karim Aoun, Meryem Lemrani, Mhaidi Idis, Aida Bouratbine, Richard Paul, Xavier Rodó

## Abstract

The development of effective Early Warning Systems (EWS) for climate-driven zoonotic diseases has been hindered by a lack of predictors with adequate lead time for effective interventions. Atmosphere-Ocean coupled phenomena present predictability beyond the atmospheric deterministic limits and therefore are potentially useful climate drivers to be integrated in mathematical models. While the El Niño-Southern Oscillation (ENSO) has been used to forecast disease dynamics in equatorial and tropical regions, there is a lack of similar applications for temperate areas, likely because of the perceived unpredictability of atmospheric systems such as the North Atlantic Oscillation (NAO). This study challenges this notion by establishing a connection between the NAO and its oceanic counterpart, the Atlantic Multidecadal Oscillation (AMO), revealing common low-frequency components that strongly modulate Cutaneous Leishmaniasis (CL) in Northern Africa. We demonstrate not only short-term couplings, such as the known NAO’s impact on seasonal rainfall, which subsequently affects CL incidence, but we also uncover a significant lagged effect of approximately three years on rainfall and four years on CL incidence. Our findings reveal a unified, multiscale mechanism that influences CL epidemiology across different time scales, underscoring the predictive skill for short and long term time frames, which should be integrated in CL forecasting models.

## Introduction

### Atmosphere-Ocean coupled phenomena and Early Warning Systems

Climate exerts a significant influence on many infectious diseases. In particular, most vector-borne diseases are influenced by prevailing weather conditions, with factors such as extrinsic incubation times, mortality and biting rates, among others, all showing sensitivity to climate variations (Caminade et al 2018). Further, the epidemiology of zoonotic diseases, known for their intricate and often delayed responses to climate, can be influenced through a multitude of complex pathways, mediated by climate-induced alterations in the landscape and vegetation, thereby impacting the population dynamics of reservoir and vector species.

Climate forcing operates across various temporal and spatial scales. Historically, infectious disease ecologists predominantly focused on examining the responses of their phenomenon of interest to local weather patterns (Stenseth et al, 2003). If strong associations emerged, this created the opportunity to make predictions days, or at a maximum weeks, ahead, depending on how slow the disease system responded to this weather forcing. As it is well known, the predictability of the atmospheric system is theoretically constrained by very short deterministic limits, making the extension of such forecasts to larger lags impossible (Tribbia et al, 1993). However, some large-scale climate processes are coupled to other more slowly-evolving systems having greater inertia (i.e. predictability), mainly in the ocean compartment. An illustrative example is the ocean-atmosphere coupled phenomenon known as the El Niño-Southern Oscillation (ENSO), which under certain conditions creates windows of predictability months (and years) in advance (Petrova et al, 2021). In fact, ENSO has already been demonstrated to be able to impact the interannual dynamics of different *tropica*l infectious diseases, allowing for the much-needed long-term predictability (Pascual et al, 2000). Under these circumstances, extended predictability beyond the deterministic limits can be observed (Chen et al, 2020).

Even when theoretically possible and of potential interest to many regions and diseases, there is a notable scarcity of operational early warning systems (EWS) designed to function on the necessary long time-scales required and providing sufficient time for effective intervention strategies. While some ENSO-based EWS exist for diseases like malaria, dengue or cholera (Pascual et al 2000, Dhiman et al 2017, Lowe et al 2011; Petrova et al., 2021; Siraj et al., 2014), there is very limited effort dedicated to zoonotic diseases, particularly in regions beyond the tropics. Importantly, when pinpointing the climate drivers behind disease transmission, merely establishing associations does not generally fit the purpose in the current highly changing climate and environmental scenarios, and it therefore becomes crucial to assemble a mechanistic explanation accounting for the observed effects in terms of epidemic outbreaks. This is especially true when dealing with delayed, nonlinear synoptic drivers whose cascading consequences can be intricate and under new conditions not covered by former data. Development of dynamical models incorporating all these important covariates becomes an optimal way to proceed, in the context of our current changing climate (Rodó et al., 2013).

### The North-Atlantic Oscillation

In temperate areas, where atmospheric-ocean couplings are weaker, apparent unpredictability has hindered the use of large-scale weather indices as disease predictors. In particular, the North Atlantic Oscillation (NAO), a dipole pattern summarized by the surface sea-level pressure difference between the Subtropical (Azores) High and the Subpolar Low (Barnston and Livezey, 1987; Luterbacher et al., 2002), is often regarded as containing no climatic memory, in terms of lead time for action. This is because it has an apparent featureless structure and displays very short memory (at most days or a few weeks) (Fernandez et al, 2003). The NAO dominates the winter climate variability over the North Atlantic and its surrounding continents (Hurrell and van Loon 1997). Despite this fact, its behavior in the low-frequency range and the dynamics behind it are not fully understood yet (Greatbatch 2000; Wanner et al. 2001; Marshall et al. 2001). Studies based on observational data have shown that extra*tropica*l sea surface temperature (SST) anomalies could force the atmospheric circulation (Czaja and Frankignoul 2002). However, in general, the ocean–atmosphere coupling can lead to a limited enhanced atmospheric low-frequency variability over the North Atlantic (for reviews see Kushnir et al. 2002; Visbeck et al. 2003; Czaja et al. 2003). An atmospheric response to midlatitude SST anomalies in combination with the oceanic gyre circulation are conceived to enhance the decadal variability of the NAO (Sutton and Allen 1997; Visbeck et al. 1998; Grötzner et al. 1998; Czaja et al. 2003; Paeth et al. 2003). In summary, this variability shows signatures at interannual timescales, as the ones explored in this study.

### Cutaneous Leishmaniasis in Northern Africa

Cutaneous leishmaniasis (CL) is one of the main parasitic vector-borne diseases affecting human beings and has an estimated annual incidence of around 1 million cases, mainly in the Americas, the Mediterranean basin, the Middle East and central Asia. CL infections in humans are generally not life-threatening and often resolve spontaneously, but they give rise to skin lesions and ulcers that can persist for months or even years, causing significant physical, psychological and social distress among affected individuals. Additionally, the disease can leave lifelong scars, leading to disability and stigma (WHO).

CL is an emerging disease in Northern Africa. CL was historically limited to several oases located in arid pre-Saharan zones, where it occurred sporadically leading occasionally to epidemics. However, since the 1980s, the geographical area of CL distribution has spread beyond its desert natural niche and an increasing number of cases are observed in new endemic foci. Currently, the disease is endemo-epidemic with thousands of cases reported every year in Libya, Algeria, Tunisia and Morocco. (Aoun et al, 2014)

In Northern Africa, CL can be caused by three *Leishmania* species: *Leishmania* (L.) *infantum, L. major*, and *L. tropica* (Aoun et al 2014). Among these, *L. major* accounts for the highest disease burden and is responsible for over 90% of the cases reported in Algeria, Tunisia, and Libya. *L. major* is a zoonotic parasite and is naturally sustained in northern Africa by a two-host cycle: rodents, such as the fat sand rat (*Psammomys obesus*) or the gerbil (*Meriones* spp). (http://www.cfsph.iastate.edu/Factsheets/pdfs/leishmaniasis.pdf) act as the main reservoirs, and *Phlebotomus* sandflies (especially *Phlebotomus papatasi*) act as vectors. Different mammals, including humans, can be infected, but most will present localized lesions and low parasitemia, thereby acting as dead-end accidental hosts and being unable to further transmit the disease. Hence, CL caused by *L. major* is referred to as Zoonotic CL (ZCL). *L. tropica*, the second most significant *Leishmania* parasite in North Africa, is on the contrary mostly sustained by anthroponotic transmission chains, thus generally transmitted between humans and sandflies without involving animal reservoirs (Saik et al 2022). As compared to Tunisia, Algeria and Libya, the epidemiological situation of CL in Morocco is characterized by a significantly higher number of cases due to *L. tropica*. This species generates an important disease burden in the area between the Atlas Mountains and the Atlantic Ocean. The incidence of *L. tropica* is limited to a few dozen cases in Tunisia, as in Algeria, and the parasite is suspected to be zoonotic with *Ctenodactylus gondii*, a rocky mountain rodent, acting as the reservoir host (Bousslimi, 2012). Rodent density, average temperature, cumulative rainfall and average relative humidity have all been shown to play a role in sustaining and increasing zoonotic CL incidence (Talmoudi et al, 2017). Furthermore, population acquired immunity may also play a role in modulating CL incidence (**Chamakh-Ayari et al, 2017**).

This study represents a significant advancement in the attempt to incorporate climate drivers typical of the northern hemisphere into epidemic-alert systems. It has the goal of deriving a multiscale EWS, by uncovering the potential for incorporating long-term predictability -up to four years in advance-for anticipating CL outbreaks in Northern Africa. While keeping with short-term signatures (i.e., the NAO link to seasonal rainfall), we here added a long-term modulation on the disease epidemiology defined by the Atlantic Multidecadal Oscillation (AMO), as we uncovered its strong link to CL dynamics in the region. To substantiate these findings, we furnished a detailed mechanistic pathway that establishes a direct link between low-frequency climate forcing and the ensuing disease dynamics. Importantly, we identified a unified, multiscale mechanism that operates both in the short term and over more extended periods.

## Data

The incidence of CL was recorded at governorate or provincial scale over a period of 17 to 23 years, using databases available at regional and national levels in Tunisia and Morocco respectively. Figure 1A shows the monthly incidence of CL caused by *L. major* in Tunisia along with the incidence of both *L. major* and *L. tropica* infections in several representative Moroccan regions. Figure 1B illustrates the mean yearly incidence of *L. major* and *L. tropica* in the selected provinces of Morocco. To ensure that the majority of CL cases are correctly attributed at *Leishmania* species level, specific provinces were selected: Errachidia, Figuig, Zagora, Jerada, and Ouarzazate being the main transmission hotspots for L. major, while Azilal, Chichaoua, Settat, and Taza being the main foci of *L. tropica*. Figure 1C displays the mean yearly incidence of CL caused by *L. major* in Tunisia at the governorate level.

**Fig. 1:**
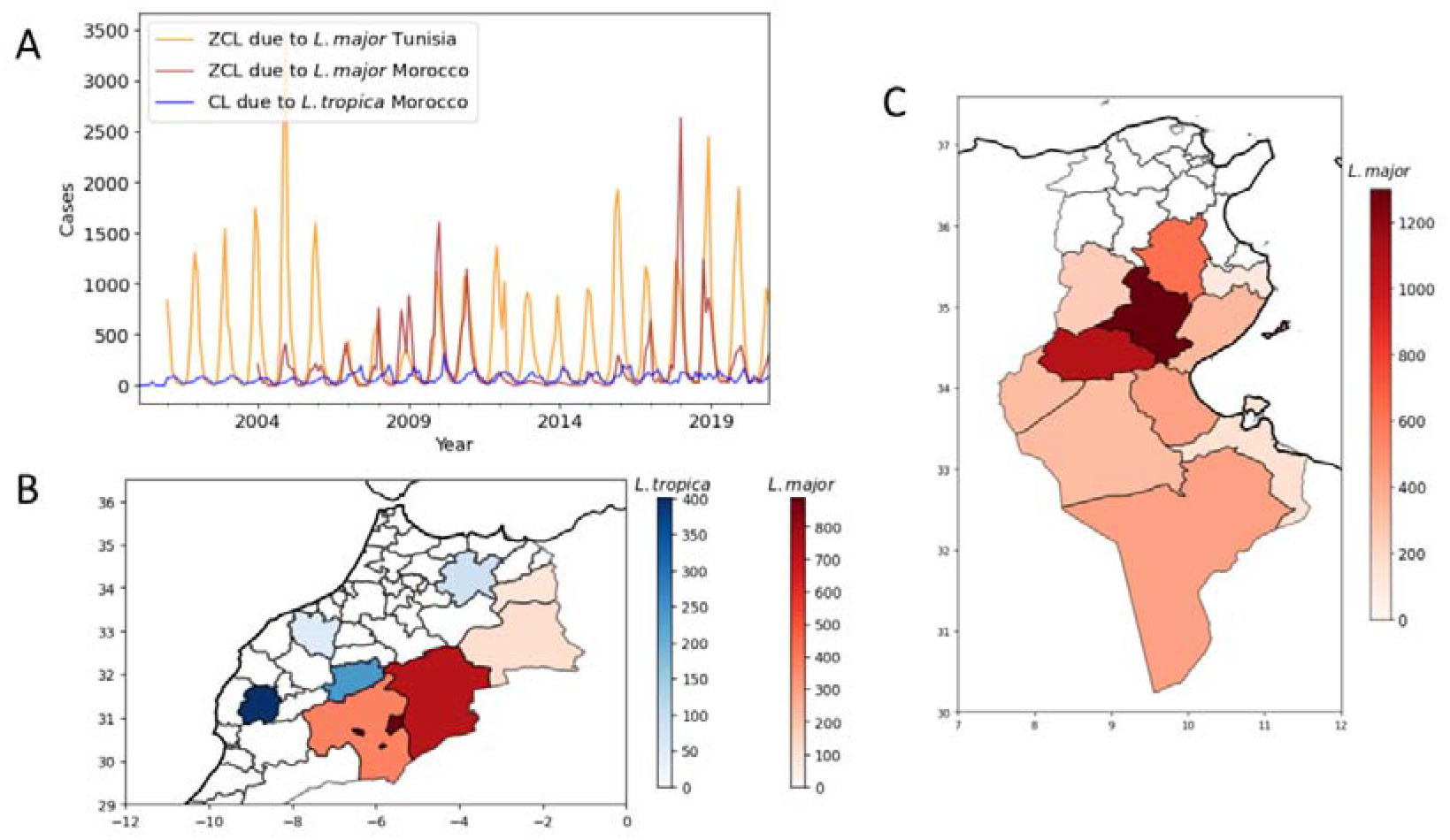
(A) Monthly incidence of *L. major* infections in Tunisia and in studied provinces of Morocco, along with the incidence of *L. tropica* infections in studied regions of Morocco. (B) Mean incidence at the province level of *L. tropica* and *L. major* in Morocco (zoomed to regions with data). (C) Mean incidence at the province level of *L. major* in Tunisia.

Reported cases of CL in North Africa show a seasonal distribution (Figure 1A). Notably, *L. major* ZCL exhibits a consistent seasonal occurrence in both Tunisia and Morocco, with case records peaking around December. This seasonal pattern of ZCL is closely related to Phlebotomus papatasi phenology in the North African region, with sandfly adult activity starting in May–June and extending to October. There is a long-lasting peak of sandfly density 3 to 3.5 months before the peak of CL incidence (Chelbi et al, 2007). The peak of *L. tropica* CL incidence around March is also likely attributed to sandfly vector phenology but also to a longer *L. tropica* incubation period compared to that of *L. major* leading to delayed onset of lesions (Aoun et al, 2022). On the other hand, Figure 1A also shows that CL incidence displays clear inter-annual fluctuations. This is particularly evident with ZCL caused by L. major, for which seasonal peaks varied from hundreds to thousands of cases according to the year (Figure 1A).

## Results

### Multiscale analysis of the NAO

Precipitation in northern Africa appears to be essentially modulated in the wet season by the North Atlantic Oscillation (NAO) and its associated pressure systems. Such pressure differences modulate the strength of the North-Atlantic westerly winds, which are responsible for the transport of moisture across the sea, and therefore impact both precipitation and temperature in extensive regions of Europe and Northern Africa (Toumi et al, 2012). CL is known to be impacted by such synoptic weather patterns, especially through modulation in precipitation. As such, we assessed whether the NAO index provided any skill that could be used in accounting in advance for CL dynamics.

Analysis of the NAO effects on ZCL in Tunisia revealed strong and clear lagged low-frequency couplings existing between NAO and ZCL incidence. Fig 2A displays both the NAO index and the ZCL incidence in Tunisia, both smoothed with a rolling mean to remove the short-term variability. When lagged around - 44 months (3.5 years), a very robust coupling emerges, which is maintained throughout the whole period. Fig 2B shows how stable the coupling is, by performing local-lagged Spearman correlations between the time series in small windows by running a Scale-Dependent Correlation (SDC) Analysis. Fig 2C shows on the spectral domain this same match between dominant periods at the low-frequency end of the spectrum, thereby reinforcing the aforementioned findings.

**Fig. 2:**
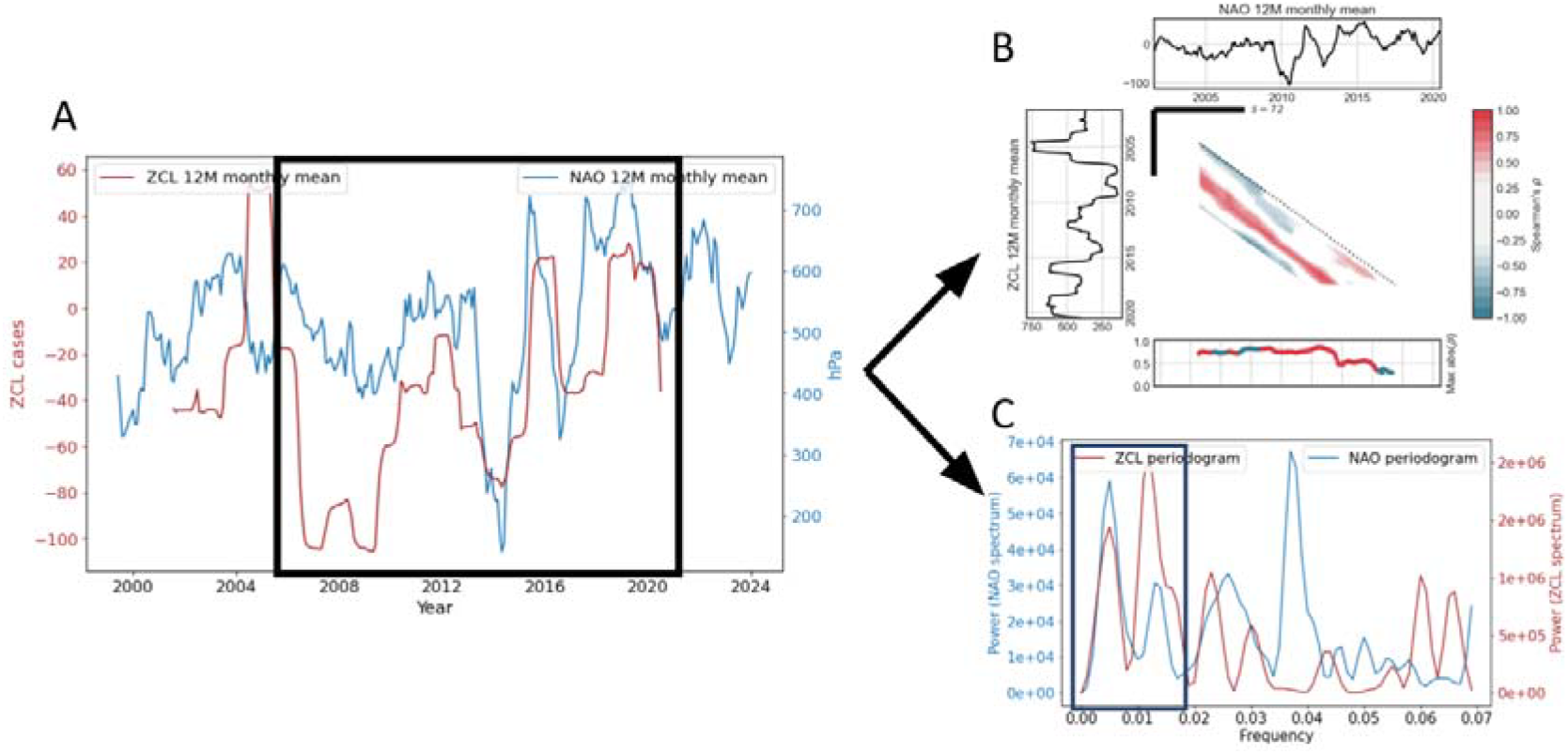
(A) ZCL incidence in Tunisia and NAO index, smoothed with a 12-M moving average and overlaid after lagging them -46 months. (B) SDC-Analysis of the two time series showing consistent strong couplings throughout the whole period. (C) Periodogram of both time series showing the match at the low-frequency end of the spectrum.

To further study the low-frequency coupling existing between the NAO index and CL, a Singular Spectrum Analysis (SSA) was applied to both the NAO and the three country-wise aggregated CL time series (*L. major* in Tunisia, *L. major* in Morocco and *L. tropica* in Morocco). Of relevance, a common low-frequency component emerged from the four aforementioned SSA decompositions with a period of around 7-10 years (Figure 3).

**Fig. 3:**
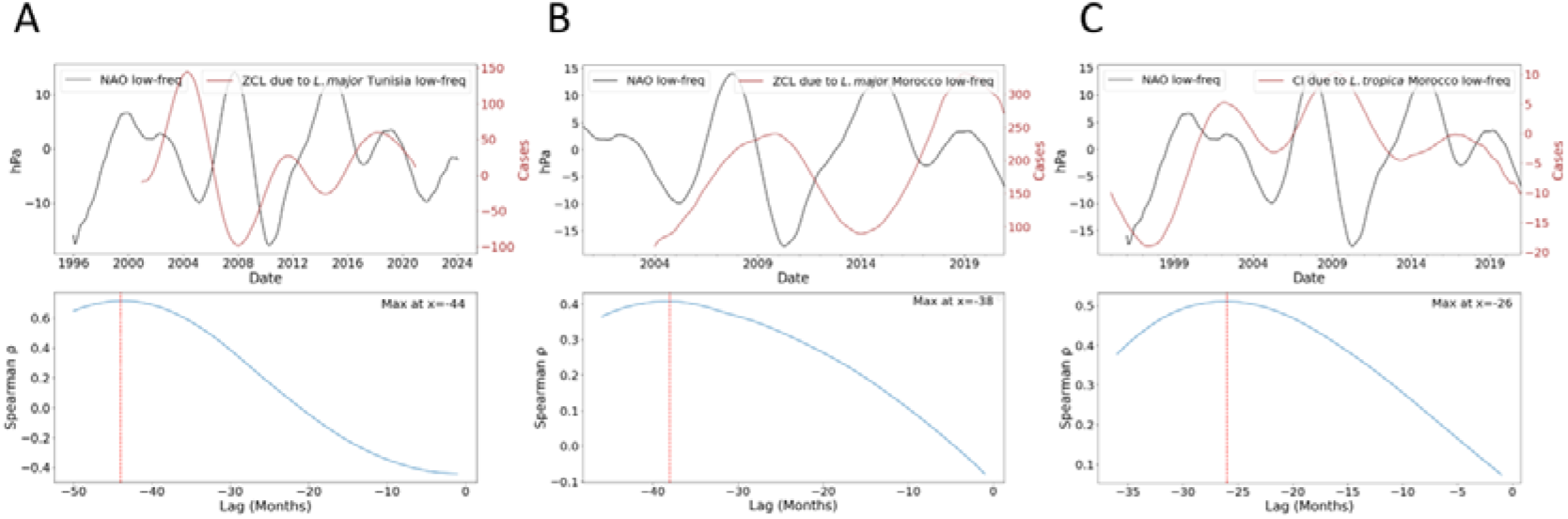
Low-frequency coupling NAO-CL. (A) For ZCL due to *L. major* in Tunisia. Up: Overlaid low-frequency components. Down: Cross-correlation analysis, showing a peak at-44 months (*p-value<0*.*05*). (B) For ZCL due to *L. major* in Morocco. Up: Overlaid low-frequency components. Down: Cross-correlation analysis, showing a peak at - 38 months (*p-value<0*.*1*; n being shorter) (C) For CL due to *L. tropica* in Morocco. Up: Overlaid low-frequency components. Down: Cross-correlation analysis, showing a peak at -26 months (*p-value<0*.*05*).

For the three CL situations, cross-correlation analyses between the low-frequency NAO and the low-frequency CL time series resulted in a unimodal function with a peak at -44 months (Tunisia L. major, *p-value<0*.*05*), -38 months (Morocco L. major, *p-value<0*.*1*; n being shorter) and -26 months (Morocco *L. tropica, p-value<0*.*05*) (Fig. 3). Similar lags, of around 3-3.5 years therefore naturally arose in Tunisia and Morocco for L. major. In parallel, a strong NAO-*L. tropica* match also emerged, despite the latter being at shorter lags.

Given the long lead times obtained, we proceeded to investigate the role and associated signatures of the Atlantic Multi-Decadal Oscillation (AMO), a coherent mode of interdecadal climate variability occurring in the North Atlantic Ocean with a fundamental estimated period of 60-80 years, based upon the average anomalies of SSTs in the North Atlantic basin.

To this end, a SSA was again computed but now for the AMO index to have it decomposed into its main modes of variability. Cross-inspection with the former SSA executed over the NAO showed common low-frequency covariating components between the two phenomena. In fact, such decompositions clearly display that the NAO-AMO coupling effectively operates synchronously across various temporal scales. The characteristic period of 7-10 years previously found in all CL time series and the NAO index, similarly shows up in the AMO (Figure 4A). Additionally, it became clear there exists a strong coherency between the 3rd component of the NAO and the 4th component of the AMO index, both showing a similar frequency peak at periods between around 2 - 3.5 years (Fig. 4). Collectively, these components elucidate approximately 17% and 7% of the AMOs and NAOs original variance, respectively.

**Fig. 4.**
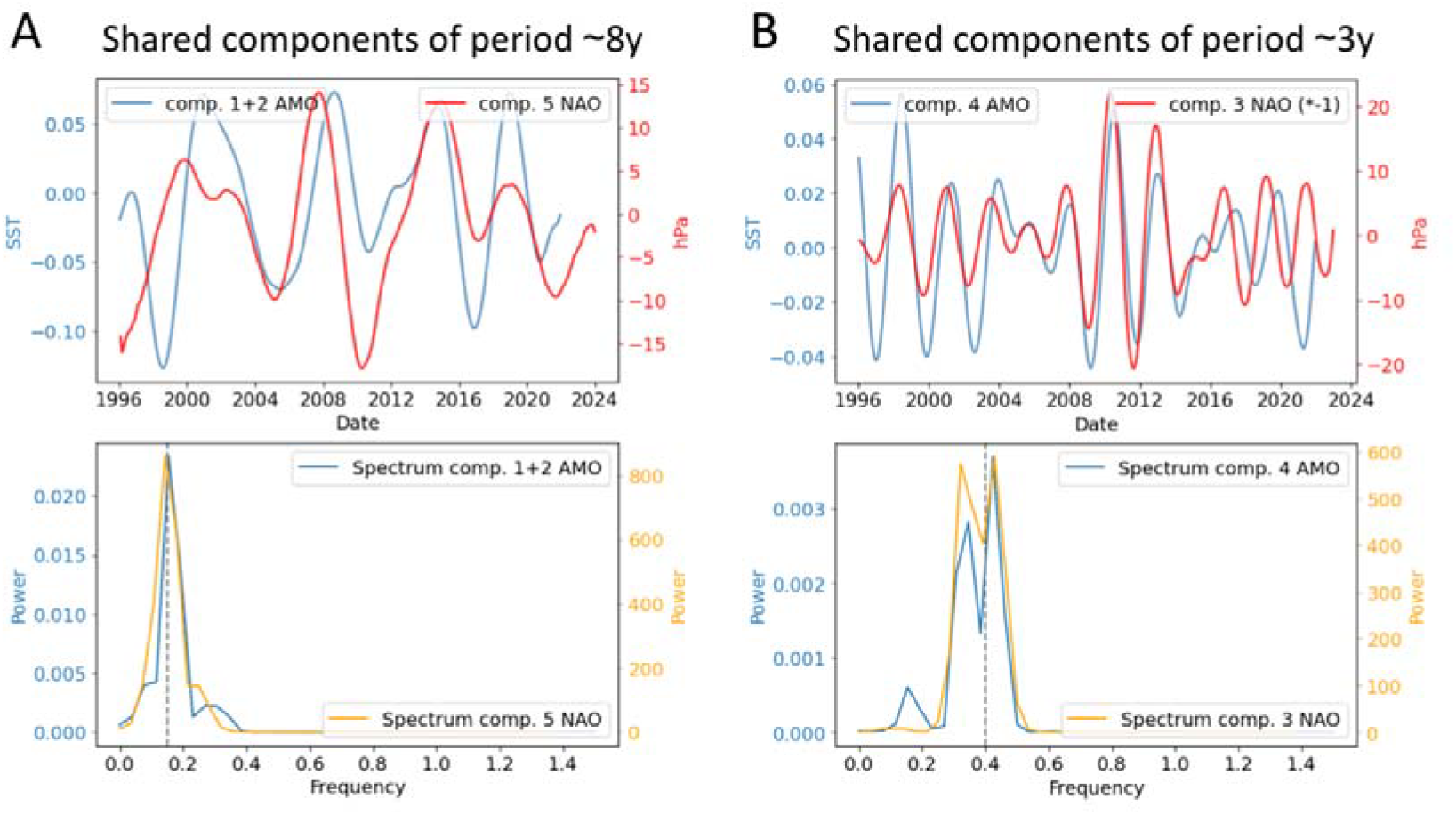
NAO-AMO coupling. (A) Shared components with a period of around 8y. (Up) Superposition of the 5th NAO component and the 1+2 AMO components (Sperman’s ρ=0.48, *p_value<0*.*05*). (Down) Overlaid Spectral decomposition of such NAO and AMO 8y components (B) Shared components of period of around 3y (Up) Superposition of the 3rd NAO component with the 4th AMO component (Spearman’s ρ=0.53, *p_value <0*.*05*). (Down) Overlaid spectral decomposition of such NAO and AMO 3y components.

The overlaid ∼8-year reconstructed components of both the leishmania parasites and the climate indices under investigation (namely, the NAO, AMO, *L. major* Tunis, *L. major* Morocco, *L. tropica* Morocco) can be seen in Fig S1.

### Short-term responses

As previously discussed, the NAO modulates the strength of the westerly winds, which are responsible for carrying moisture across the Atlantic, and therefore they seasonally impact both precipitation and temperature in Europe and Northern Africa. It is furthermore surprising to find a large lagged effect given that the NAO instead has an instant effect on precipitation and quickly loses predictability (Wunsch, 1999). Indeed, at these very short time scales ZCL in Tunisia is seen to be strongly associated with synoptic weather events displaying a dipole linked to the NAO phenomenon through the Açores pole (Fig S3), despite being also partly related to the Scandinavian Mode (Wang and Tang, 2020). Its associated skill can be traced back to a maximum of 6 months in the form of leading sea-level pressure anomalies (Fig S3).

To further delve into the climatic forcing mechanisms operating on ZCL dynamics, we zoomed in and regionalized the analyses in Tunisia. We identified an expected significant short-term coupling between the NAO and precipitation raw values (not shown), which also showed up after applying a 12-month smoothing filter (Fig 5A). However, apart from the short-term coupling, the associations at longer lags previously identified similarly showed up (Fig 5A).

**Fig. 5:**
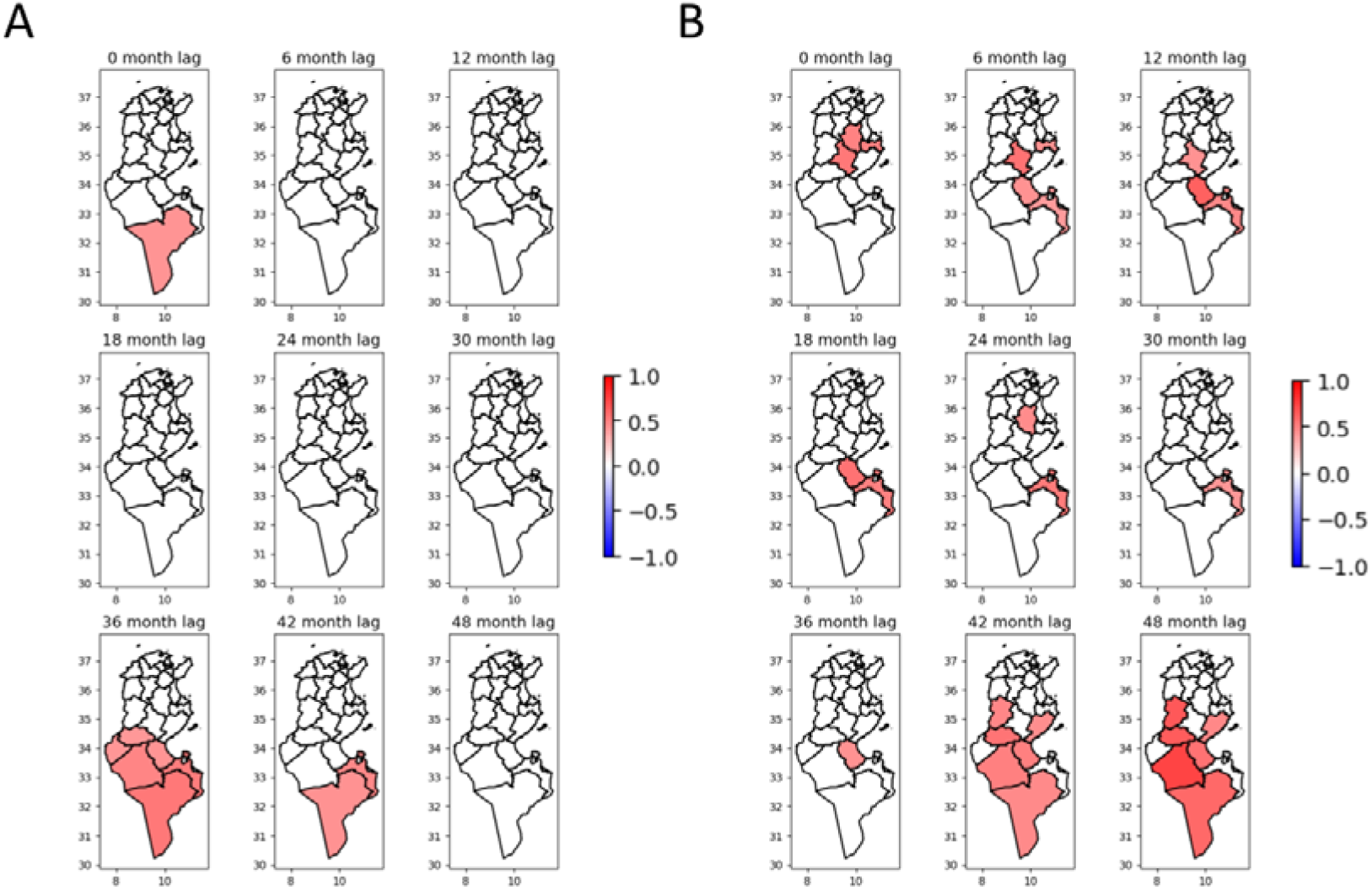
(A) Significant full series correlations (*p-value < 0*.*05*) of NAO vs. Rainfall (smoothed with 12-mo moving windows) in Tunisia at the province level from 2007 to 2021. (B) Significant correlations (*p-value < 0*.*05*) NAO vs ZCL (smoothed with a 12-mo moving average) in Tunisia at the provincial level for the period 2007-2021

In the previous analysis a maximum low-frequency NAO-ZCL coupling was found for Tunisia with a 44 months delay (Fig 2A). We now obtained a maximum NAO-Precipitation coupling located at 36 months delay, therefore around 1 year before that obtained for the NAO-ZCL, thereby reinforcing our previous findings and showcasing how precipitation mediates in the cascade effects of NAO on ZCL in Tunisia.

We proceeded to perform the same analyses but, now instead, directly between the NAO and ZCL. In a higher spatial resolution domain, we again obtained evidence regarding the existence of a strong NAO-ZCL coupling at around -44 months lag, manifesting especially in the middle and southern part of Tunisia, where we also saw the best couplings with precipitation. (Fig 5A, 5B)

The relationship between NAO and precipitation in Morocco is less direct and evident compared to the situation in Tunisia (Figs S5, S6).

Local precipitation impacts ZCL short-frequency dynamics in both countries (Fig. 6). At the Governorate level / Wilaya level, ZCL and precipitation tend to show a maximum coupling at around a year lag. Interestingly, the normalized difference vegetation index (NDVI), known to respond tightly to rainfall in these types of climates, displayed higher correlations with ZCL at slightly shorter lags. Figure 9 shows a noticeable example of such coupling for Errachidia (Morocco) and Gafsa (Tunisia).

**Fig. 6.**
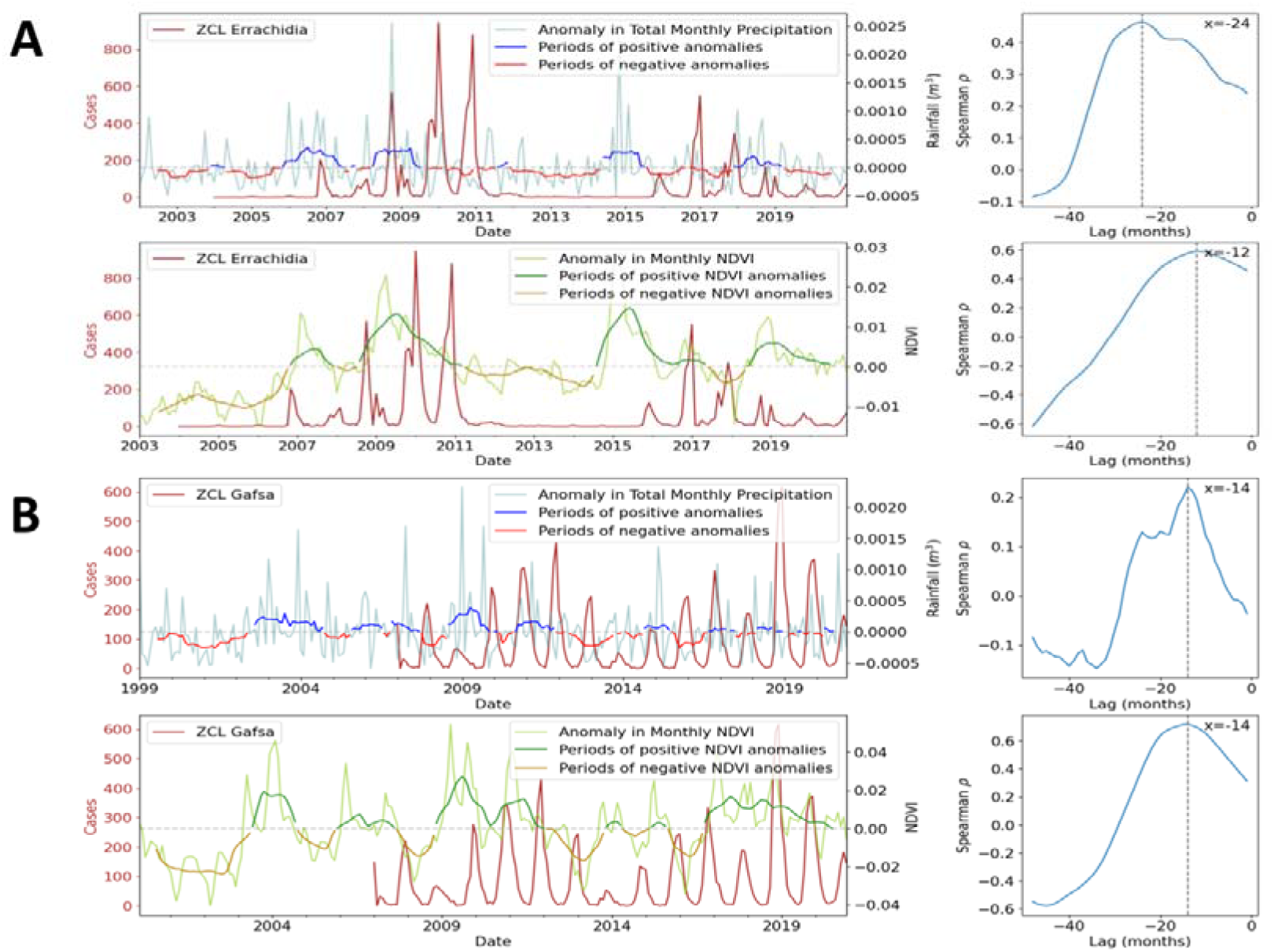
ZCL-Rainfall couplings (up) and ZCL-NDVI couplings (down) -and cross-correlation analyses (CCF), right-for (A) Errachidia (Morocco) and (B) Gafsa (Tunisia). All original rainfall, NDVI and ZCL time series are shown (in light blue, light green and brown). The graphs also display the result of 12-mo moving filter on the climatic and environmental time series, which serve to illustrate periods of positive anomalies (in blue for precipitation and green for NDVI) and negative anomalies (in red for precipitation and yellow for NDVI). CCFs are performed after applying a 12-mo moving filter to all time series to remove short term variability. The correlation peaks for the cross-correlations from Errachidia all pass a significance test at alpha=0.05. For Gafsa, and mainly because the timeseries is shorter, the cross-correlation peak with rainfall doesn’t pass a test (p_value <0.1) but it does with NDVI.

## Discussion

Very few studies have attempted to investigate the inter-annual modulation of CL and associate it to external forcings. One seminal study identified 3y cycles in CL data from Costa Rica and hypothesized them to be connected to the El Niño Southern Oscillation (ENSO) phenomenon (Chaves et al, 2006). A shared frequency peak was identified between CL incidence and the ENSO MEI index, but the study did not provide a detailed mechanistic explanation of how the atmosphere-ocean coupling takes place from the source region in the equatorial SST and the local CL dynamics in Costa Rica. As such, the exact mechanistic cascading pathway underlying the findings remained elusive.

In this study, instead, we proved that despite the distance between Morocco and Tunisia, and the large known differences between *L. major* and *L. tropica* epidemiological cycles, all the studied different CL disease situations in North-Africa show strong low-frequency components being shared with the NAO. In fact, such couplings take place at very similar lags for the two *L. major* situations, but at a different lag for *L. tropica*. This apparent discrepancy should not be surprising and reinforces these findings. *L. tropica* represents a non-zoonotic form of CL, that does not appear to operate through an animal reservoir. Therefore, we hypothesize that it presumably responds faster to the same extrinsic climatic stressor, which would explain the shorter lags found. Overall, we inferred that CL inter-annual dynamics are governed by a large-scale climate synoptic pattern, which likely operates along with population acquired immunity to determine the final low-frequency patterns of the CL time series (Chamakh-Ayari et al, 2017).

Overall, we identified two shared components between the NAO and the AMO illustrating that both are showing responses to the same climatic process. Therefore, from a mechanistic perspective, a unique unified mechanism can explain both the multiscale short-term and long-term relationships between climate and ZCL epidemiology in Northern Africa.

Regarding the exact pathways by which such large-scale indices locally affect transmission, we identified a strong modulation of precipitation in Tunisia, both instantaneous and at 3 years lag. On the latter it is important to say that such a 3-year lag is around one year before the maximum couplings appear with respect to the disease (which manifest around a 4-year lag). This one-year difference between the cascading effects of precipitation on the disease (i.e., before effects show up in ZCL incidence) was also independently identified in Fig 9, thereby reinforcing our findings. Furthermore, by integrating data from vegetation, which strongly couples to ZCL incidence, we found evidence that supports the notion that vegetation plays a key role in sustaining and increasing zoonotic CL incidence. These results are in line with those of other studies showing that the incidence of ZCL in central Tunisia increased significantly when there was an increase in rainfall lagged by 12 to 14 months (Toumi et al, 2012). They are also in accordance with results of other studies that found that the occurrence of ZCL is related to rodents’ density but lagged 2 months (Talmoudi et al, 2017). In fact, higher rainfall is expected to result in increased density of vegetation, particularly for chenopods, a halophytic plant that constitutes the exclusive food source of *P. obesus*. Presumably, following a high density of *P. obesus*, the source of *L. major* transmissible from the rodents to blood-feeding female sand flies could lead to a higher probability of transmission to humans over the next season.

In Morocco, the NAO did not so clearly appear to modulate precipitation, especially when aggregating data from the whole country. Morocco’s unique and complex geographical position, being in a fringe topographic region separated by the Atlas Mountains, may be behind the fainter signature -albeit also significant-obtained with CL. This mountain range is responsible for the distinct manifestation of precipitation patterns on either side, ultimately being influenced by diverging interactions with the Atlantic moist winds and steep topography. The nonlinearities in the observed Morocco’s ZCL response to the NAO could also be due to these more complex, multifaceted pathways by which the NAO modulates climate in northern Africa.

## Conclusions

The AMO low-frequency oceanic pattern in the North Atlantic modulates the low-frequency dynamics of ZCL epidemiology in North Africa. Its fundamental role can explicitly be traced to the cascading effects connecting the long-term SST changes in the North Atlantic to the ZCL dynamics. In particular, we could show how some of the main modes of variability of the AMO are transferred via the NAO. Physically, this means that variations in the SST anomalies of the Atlantic Ocean may operate in the form of large-scale changes in synoptic weather conditions subsequently affecting the moist westerly winds patterns over the Atlantic. This in turn creates changes in precipitation across North Africa that can be traced to ultimately affect CL incidence through cascading effects in vegetation.

A multi-scale prediction system such as the one described in this study, that can operate at both long (roughly 4 years) and short (6 months) having the NAO and the AMO as the sources of climatic memory, therefore predictability, has never been built before. Importantly our results identify a unique multiscale mechanism accounting for both short-term and long-term dynamics, that could be promptly integrated to provide an alert system for the different types of cutaneous leishmaniasis in the region.

## Methods

Scale-Dependent Correlation (SDC) analysis is an optimal method for identifying dynamical couplings in short and noisy time series (Rodo et al, 2006). In general, Spearman correlations between incidence and a meteorological time series assess whether there is a monotonic relation between the variables. SDC analysis was specifically developed to study transitory associations that are local in time at a specified temporal scale corresponding to the size of the time intervals considered (s). The two-way implementation (TW-SDC) is a bivariate method that computes non-parametric Spearman rank correlations between two time series, for different pairs of time intervals along these series. Different window sizes (s) can be used to examine increasingly finer temporal resolution. Significance is assessed with a non-parametric randomization test. For the baseline test, SDC calculates Spearman correlations (at α⍰=⍰0.05) between two white-noise time series at each fragment size s for a non-parametric permutation test. The indices of the series are randomly re-ordered, breaking their temporal shape. This permutation test enables a first estimate of the probability of finding significant spurious correlations, and it can thus be used as a non-parametric significance test for pairs of any length for the time series of interest. (The code in Python for Scale-Dependent Correlation Analysis (SDC) developed by X.R. can be found at https://github.com/AlFontal/sdcpy.)

Singular spectrum analysis (SSA) (Ghil et al 2002, Vautard et al 1992) was applied to separate different orthogonal components in both the CL time series and the climate ones. SSA involves the spectral decomposition (eigenvalues and corresponding eigenvectors) of a covariance matrix obtained by lagging the time series data for a prescribed number of lags M called the embedding dimension. There are two crucial steps in this analysis for which there are no formal results but useful rules of thumb: one is the choice of M; the other is the grouping of the eigenvectors to define the specific major components and reconstruct them. Typically, the grouping of the eigencomponents is based on the similarity and magnitudes of the eigenvalues, their power (variance of the data they account for), and the peak frequency of resulting reconstructed components (RC). For the selection of the embedding dimension one general strategy is to choose it so that at least one period of the lowest frequency component of interest can be identified, that is M⍰>⍰fs/fr, where fs is the sampling rate and fr is the minimum frequency. Another strategy is that M be large enough so that the M-lagged vector incorporates the temporal scale of the time series that is of interest. The larger the M, the more detailed the resulting decomposition of the signal. In particular, the most detailed decomposition is achieved when the embedding dimension is approximately equal to half of the total signal length. A compromise must be reached, however, as a large M implies increased computation, and too large a value may produce a mixing of components. SSA, given its data-adaptive nature and orthogonal decomposition approach, can be used as an alternative smoothing method to moving averages, which is more flexible and robust, and also deals better with non-stationary noisy time series (Ghil et al., 2002).

Rainfall in semi-arid regions typically exhibits a lot of stochasticity. We applied moving averages in some situations to extract lower-frequency components. In such instances, and also in the analysis of low-frequency components, given that both series become more autocorrelated and their number of degrees of freedom decreases, we calculated p-values by correcting by the effective degrees of freedom. In particular, we employed a bootstrap approach to assess their significance. To compute the distribution of resulting correlation coefficients from which we determined significance, we repeatedly randomized the smoothed time series while keeping original autocorrelation.

To study responses in vegetation to rainfall variability ultimately impacting CL, we used the normalized difference vegetation index (NDVI). NDVI is a remote sensing measurement used to assess the density and health of vegetation. It calculates the difference between near-infrared (which vegetation strongly reflects) and red light (which vegetation absorbs) from satellite or aerial imagery, providing a numerical indicator that can be used to monitor plant growth, vegetation cover, and biomass production.

## Data Availability

All data produced in the present study are available upon request to the authors

## Acknowledgements

The authors acknowledge Drs Latifa Maazaoui and Kaouther Hrabech from the Tunisian Ministry of Health for their precious help in the collection of CL data.

A.SJ was supported by a fellowship from “la Caixa” Foundation, Spain (ID 100010434, fellowship code LCF/BQ/DR19/11740017). A. SJ and X.R. acknowledge the support from the grant CEX2018-000806-S funded by MCIN/AEI/10.13039/501100011033 and support from the Generalitat de Catalunya through the CERCA Program.

